# Effects of blood pressure and arterial stiffness on retinal neurodegeneration: Cross-sectional and longitudinal evidence from UK Biobank and Chinese cohorts

**DOI:** 10.1101/2022.09.27.22280405

**Authors:** Yining Huang, Yixiong Yuan, Yujie Wang, Ziwen Hui, Xianwen Shang, Yanping Chen, Siran Zhang, Huan Liao, Yifan Chen, Mingguang He, Zhuoting Zhu, Wei Wang

**Affiliations:** Nanshan School, Guangzhou Medical University, Xinzao Road, Panyu District, Guangzhou 511436, China; Department of Ophthalmology, Shanghai General Hospital, Shanghai Jiao Tong University School of Medicine, Shanghai, China; Zhongshan school of medicine, Sun Yat-sen University, Guangzhou, China; Centre for Eye Research Australia, Royal Victorian Eye and Ear Hospital, Melbourne, Australia; Department of Ophthalmology, Guangdong Academy of Medical Sciences, Guangdong Provincial People’s Hospital, Guangzhou, China; State Key Laboratory of Ophthalmology, Zhongshan Ophthalmic Center, Sun Yat-sen University, Guangdong Provincial Key Laboratory of Ophthalmology and Visual Science, Guangdong Provincial Clinical Research Center for Ocular Diseases, Guangzhou, China; Epigenetics and Neural Plasticity Laboratory, Florey Institute of Neuroscience and Mental Health, University of Melbourne; John Radcliffe Hospital, Oxford University Hospitals NHS Foundation Trust, Oxford, UK

**Author notes:** Co-first authors (Yining Huang, Yixiong Yuan). Co-corresponding authors. **Corresponding authors:** Wei Wang, MD PhD, Zhongshan Ophthalmic Center, State Key Laboratory of Ophthalmology, Sun Yat-sen University, Guangzhou, China., Zhuoting Zhu MD PhD, NHMRC Emerging Leadership Fellow, Ophthalmic Epidemiology, Centre for Eye Research Australia, University of Melbourne. Level 7, 32 Gisborne Street, East Melbourne, VIC 3002, Australia., |, Mingguang He, MD PhD FRANZCO, NHMRC Leadership Fellow, Professor of Ophthalmic Epidemiology, Centre for Eye Research Australia, University of Melbourne, Level 7, 32 Gisborne Street, VIC 3002, Australia., |.

## Abstract

**Background:** Retinal neurodegeneration, an easily accessible biomarker of dementia risk, is exacerbated by age and hypertension; however, the relative roles of systolic and diastolic blood pressure (SBP and DPB) remain unclear. This study aimed to determine the cross-sectional and longitudinal associations between BP and atherosclerosis levels along with the different retinal neurodegeneration parameters.

**Methods:** This study used cross-sectional data from the United Kingdom (UK) BioBank (UKB) and longitudinal data from the Chinese Ocular Imaging Project (COIP). The macular ganglion cell-inner plexiform layer thickness (mGCIPLT) and macular retinal nerve fiber layer thickness (mRNFLT) were measured using spectral domain optical coherence tomography imaging. Swept-source optical coherence tomography was performed at each follow-up visit to obtain the longitudinal trajectory of the mGCIPLT and peripapillary RNFLT (pRNFLT) in the COIP cohort. Multivariable linear models were used to analyze the cross-sectional and longitudinal associations between BP metrics and retinal measurements.

**Results:** In a cross-sectional analysis of 22,801 participants from the UKB, thinner mGCIPLT was related to older age (per 10 years, β = −0.274, 95% confidence interval (CI): −0.358 to −0.189, p < 0.001), female sex (female vs male, β = −0.897, 95% CI: −1.031 to −0.763, p = 0.000), higher SBP (per 10 mmHg increase, β = −0.085, 95% CI: −0.125 to −0.045, p = 0.000), and higher DBP (per 10 mmHg increase, β = −0.105, 95% CI: −0.174 to −0.036, p = 0.003), and was significantly associated with higher mean arterial pressure (MAP per 10 mmHg increase, β = −0.116, 95% CI: −0.176, −0.056, p = 0.000) and higher mean pulse pressure (MPP per 10 mmHg increase, β = −0.099, 95% CI: −0.155, −0.043, p = 0.001). In a longitudinal analysis of 2,012 eligible COIP participants, higher levels of baseline SBP, DBP, MAP, and MPP were associated with faster thinning in mGCIPLT and pRNFLT (all p < 0.001). The strongest association was with MAP, which produced an effect on mGCIPLT (β = −0.118, 95% CI: −0.175 to −0.061, p < 0.001) per 10 mmHg increase, comparable to a 5-year increase in age (β = −0.210, 95% CI: −0.282 to −0.138, p < 0.001). The results of the analysis of mRNFL and pRNFL were consistent with those of mGCIPLT.

**Conclusion:** BP levels were independently and consistently associated with various retinal neurodegenerative exacerbations, both cross-sectionally and longitudinally, regardless of the race and disease status. BP plays a key role in neurodegeneration, and long-term prevention in the population requires the control of BP levels.

**Graphic abstract:** 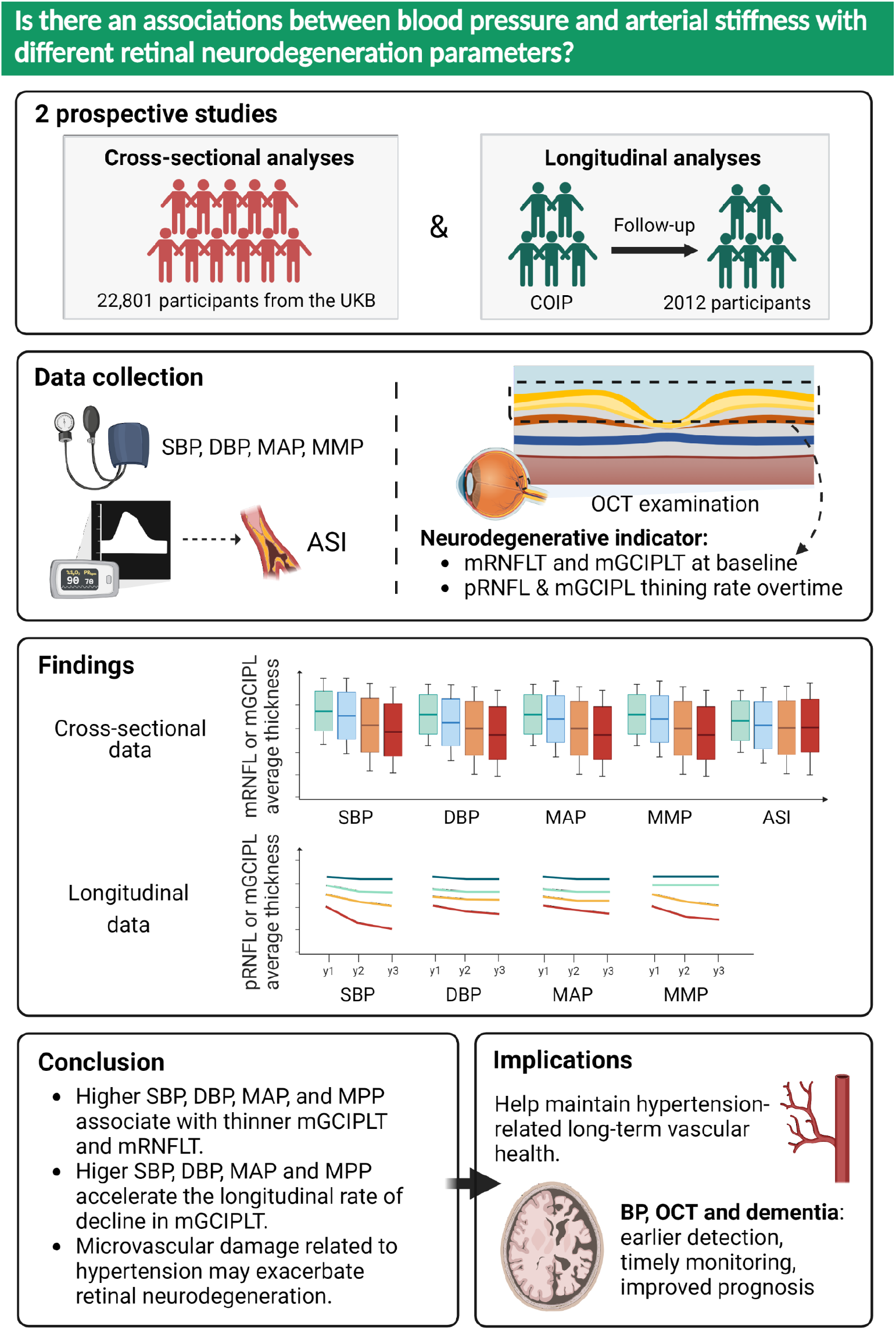

## Introduction

Dementia is one of the leading causes of morbidity and mortality in older adults worldwide.^1–4^ Retinal neurodegeneration is a comprehensive, early window into the risk of dementia.^5^ Retinal optical coherence tomography (OCT) imaging achieves unprecedented resolution; in Alzheimer’s disease (AD) patients, OCT can detect progressive thinning of the retinal nerve fiber layer (RNFL) and the ganglion cell-inner plexiform layer (GCIPL).^6^ There is currently a sudden increase in the use of OCT in the community and hospitals, such as in all optometric branches of Specsavers, the largest optometric company in the United Kingdom (UK), which is equipped with OCT, providing a noninvasive approach to dementia risk stratification and early preventative treatment.

Hypertension is another major global disease that has a significant impact on the neurological structure and function.^7^ Our previous study found that hypertension can cause a decrease in brain volume, which increases the risk of dementia, and that the earlier the age of diagnosis, the more damage hypertension does to the brain and the greater the risk of dementia.^8^ Hypertension is a strongly treatable risk factor; meta-analytical evidence suggests that antihypertensive medications reduce the risk of dementia, and conclusive evidence shows that lowering the blood pressure (BP) reduces the risk of dementia. However, data on the patient group that will most likely benefit from such treatment are lacking.^9–12^ The critical period of exposure to elevated BP, and therefore the optimal period of treatment, remains unknown.

The relationship between RNFL thickness (RNFLT), GCIPL thickness (GCIPLT), and BP has received much attention as a surrogate indicator of dementia risk. Several studies have reported thinner RNFLT and GCIPLT in patients with hypertension lasting more than 5 years compared with the healthy population.^13,14^ In individuals with type 2 diabetes mellitus (DM), the peripapillary RNFLT (pRNFLT) was lower in those with hypertension as a comorbidity than in those with DM alone, and the hypertension duration was significantly associated with pRNFLT.^15^ However, other studies have shown that neither systolic blood pressure (SBP) nor diastolic blood pressure (DPB) has an effect on the rate of retinal neurodegeneration.^16,17^ Moreover, an inverted U-shaped association was found between BP status and GCIPLT and RNFLT, and that both hypotension and hypertension were associated with thinning of the inner retinal layers.^18^ However, most of these studies were cross-sectional in nature with small samples, and only a few longitudinal studies were conducted. To the best of our knowledge, no study has specifically analyzed the association between different BP metrics and longitudinal changes in RNFLT and GCIPLT.

The UK BioBank (UKB), a population-based cohort of more than 500,000 people, is the largest OCT dataset available and offers a unique opportunity to clarify the association between BP and retinal neurodegeneration; however, it lacks data on the longitudinal changes in OCT and data on Chinese ethnicity. Therefore, this study aimed to perform a cross-sectional analysis of the UKB and a longitudinal analysis of the Chinese cohort to clarify the effect of different BP metrics on RNFLT and GCIPLT.

## Methods

### Design and participants

The overall design of this study is illustrated in **Figure 1**. Data from the UK BioBank (UKB) and Chinese Ocular Imaging Project (COIP) cohorts were used in this study. A cross-sectional relationship analysis between BP, RNFLT, and GCIPLT was performed on the UKB data, while a longitudinal relationship analysis was performed on the COIP cohort. The UKB and COIP were approved by the NHS North West Multicenter Research Ethics Committee (application no. 62491) and the Institute Ethics Committee of Zhongshan Ophthalmic Center (no. 2017KYPJ094), respectively. All participants provided written informed consent, and the study was performed in accordance with the Declaration of Helsinki. The specific methods used in both studies have been reported elsewhere and are briefly described below.

**Figure 1.**
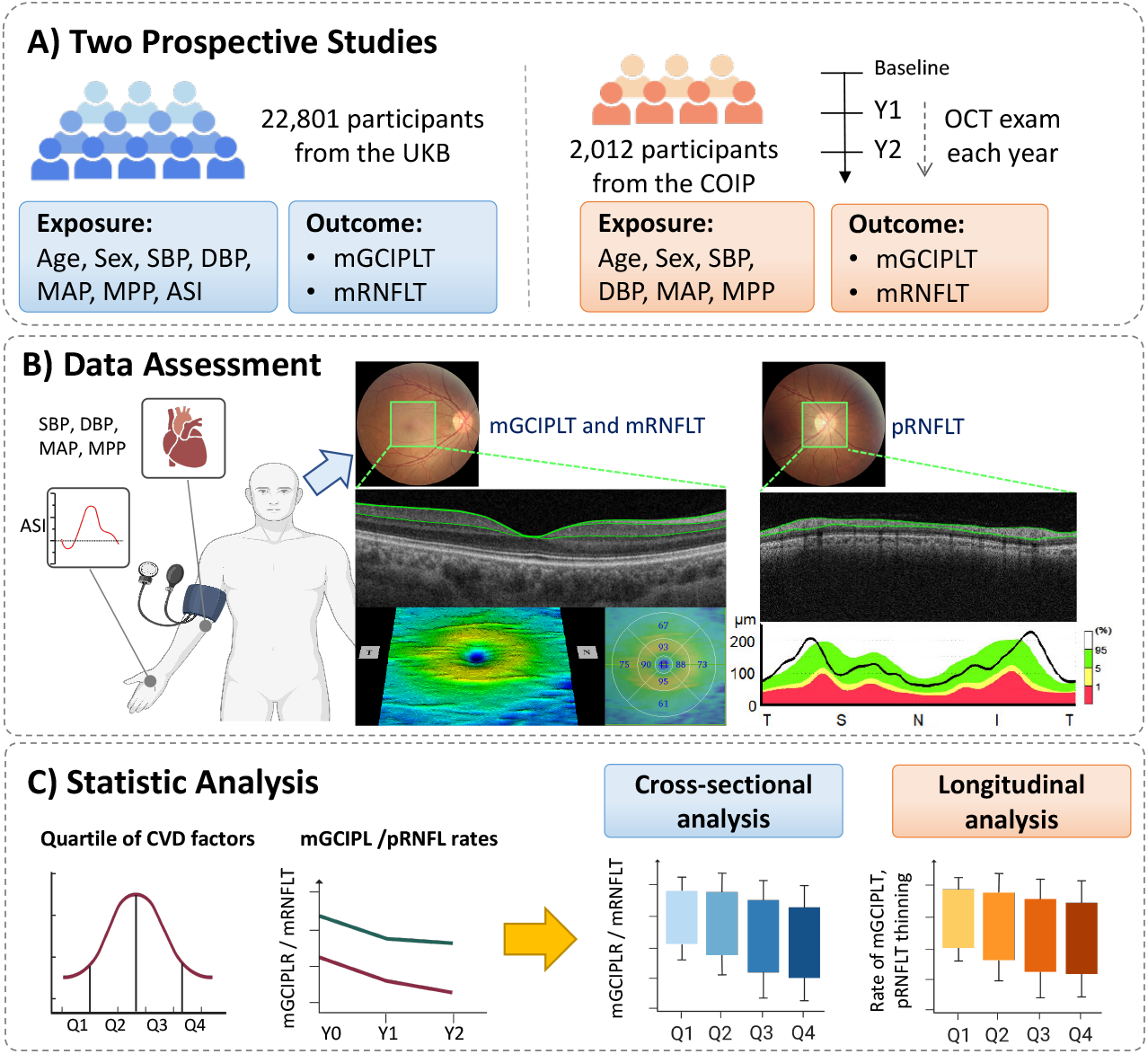
Overall workflow of the study.

#### UKB

From 2006 to 2010, the UKB study recruited 502,639 participants, aged 40–69 years, from the general population across the UK; used questionnaires; and undertook trials at 22 testing centers in England, Scotland, and Wales. Those with qualifying OCT and BP data were considered to meet the inclusion criteria. The exclusion criteria were as follows: high refractive error, visual acuity of <20/200, intraocular pressure (IOP) of ≤5 or ≥22 mmHg, dementia, Parkinson’s disease, obstructive sleep apnea syndrome, multiple sclerosis, age-related macular degeneration, glaucoma, diabetic eye disease, and missing data on important variables.

#### COIP

Between November 2017 and December 2018, the COIP cohort consecutively recruited patients with DM and healthy individuals from the Guangzhou community for comprehensive ophthalmic and systemic examinations. The absence of retinopathy and optic neuropathy was confirmed using Early Treatment Diabetic Retinopathy Study (ETDRS) 7-field fundus photography and OCT. The exclusion criteria were similar to those used for screening the UKB population: major systemic pathologies other than diabetes and hypertension, a history of intraocular surgery or laser treatment, and a history or evidence of retinal or optic nerve disease. The study participants underwent standard annual systemic and ocular examinations.

### Retinal OCT imaging to assess for neurodegeneration

#### UKB

A total of 67,321 patients from six testing centers underwent macular retinal imaging using a spectral domain OCT device (Topcon 3D OCT-1000 Mark II), which has an axial resolution of 5 μm and a lateral resolution of 15 μm. OCT imaging was performed without pupil dilation in a dark room using a three-dimensional (3D) 6 × 6 mm^2^ macular volume scan mode (512 A-scans per B-scan, 128 horizontal B-scans in a raster pattern). The Topcon advanced boundary segmentation algorithm was used to segment the boundaries of the retinal layers. RNFLT was defined as the distance from the inner boundary membrane (ILM) to the nerve fiber layer, while GCIPLT was defined as the distance from the RNFL/GCIPL junction to the ILM/inner nuclear layer. The average thickness of the macular RNFL (mRNFLT) and macular GCIPL (mGCIPLT) in the ETDRS regions were obtained. Only high-quality OCT image data were used in this study, and data with the following conditions were excluded: signal strength of <45, poor centration, and segmentation errors.

#### COIP

After the participants’ pupils were dilated, macular and optic disc imaging was performed by the same examiners using a commercially available swept-source OCT device (DRI OCT Triton; Topcon, Japan). Each OCT scan was performed with an internal fixator, and fixation was monitored using the instrument’s built-in fundus camera. The retina was imaged using a 3D macula cube 7 × 7-mm scan mode centered on the central macular recess with a scan density of 512 A-scans × 512 B-scans. A 3.4-m ring-scan model centered on the optic disc was used. Images with a quality score of <60, segmentation errors, presence of artifacts, dark areas due to blinking, eccentric scans, and missing scan lines were excluded. The pRNFLT and mGCIPLT measurements in the average, upper, lower, nasal, and temporal quadrants were obtained using a built-in automated algorithm.

### Blood pressure and arterial stiffness index measures

Two BP and heart rate measurements were performed using an Omron 705 IT electronic BP monitor, with at least 1-min interval between measurements; the average of the measurements was used for analysis. The patient’s history of BP medication treatment was assessed to confirm the use of antihypertensive drugs. Hypertension was defined as an SBP of ≥140 mmHg, a DPB of ≥90 mmHg, or self-reported use of antihypertensive medications. The mean arterial pressure (MAP) was calculated as [(2 × diastolic pressure) + systolic pressure]/3. The mean pulse pressure (MPP) was calculated as the SBP minus the DPB. Arterial stiffness was calculated using pulse waveforms obtained from noninvasive measurements of the finger using an infrared sensor (Pulse Trace PCA2, CareFusion). The participant’s height in meters was divided by the time when the pulse waveform peaks to calculate the arterial stiffness index (ASI) in meters per second. In the COIP, the BP parameters were measured using the same methodology used in the UKB, but the ASI data were not available owing to the absence of infrared sensor equipment.

### Assessment of systemic and ocular covariates

#### UKB

Data on age, sex, and other sociodemographic characteristics were collected using questionnaires. The patients’ history of myocardial infarction, stroke, and diabetes was determined by self-report. Height was measured using a Seca 202 height meter and expressed in centimeters. Body weight was measured to the nearest 0.1 kg using a Tanita BC-418 body composition analyzer. Body mass index (BMI) was calculated as body weight in kilograms divided by height in meters squared. The history of cardiovascular disease and DM was self-reported. The serum biomarkers were measured using a clinical chemistry analyzer. The ocular parameters included visual acuity and IOP (ocular response analyzer, Reichert, Depew, NJ, USA).

#### COIP

All participants completed a comprehensive questionnaire and underwent annual eye examinations. Data on age, sex, T2DM duration, and history of systemic disease were collected using a standardized questionnaire. SBP, DBP, height, and weight were measured using standardized procedures. The venous blood samples and clean intermediate urine samples were collected for the determination of serum total cholesterol, high-density lipoprotein cholesterol, low-density lipoprotein cholesterol, triglyceride (TG), glycated hemoglobin (HbA1c), and urine microalbumin levels. Other eye examinations included slit lamp examination (BQ-900, Haag-Streit, Köniz, Switzerland) and indirect ophthalmoscopy for fundus examination. IOP was measured using a non-contact tonometer (CT-1 Tonometer, Topcon Ltd., Topcon). Visual acuity and best-corrected visual acuity were measured at a distance of 4 m using an ETDRS logMAR E visual acuity meter (Precision Vision, Villa Park, Illinois, USA). The axial length and central corneal thickness were measured using a Lenstar LS900 biometer (HAAG-Streit AG, Köniz, Switzerland). Myopia and astigmatism were assessed using an automated optometer (Topcon KR8800; Topcon Corporation, Tokyo, Japan).

### Statistical analyses

Only data from the right eye were included in the statistical analysis. The continuous variables were presented as mean ± standard deviation (SD), while the categorical variables were presented as numbers (percentages). All variables were tested for normality; if normality was not satisfied, the variables were transformed into normality, or nonparametric tests were used.

For the cross-sectional analysis of the UKB, BP was set as a continuous variable, and statistical analysis was performed using the same method as previously reported. Linear models were used to investigate the relationships between age, sex, SBP, DBP, MAP, MPP, ASI, mRNFLT, and mGCIPLT. Model 1 was adjusted for age, sex, race, and testing center; model 2 was additionally adjusted for smoking, BMI, alcohol, Townsend deprivation index score, and education level. Model 3 extended model 2 by further adjusting for HbA1c, total cholesterol, and TG levels based on model 2, and excluded patients with a history of DM, hypertension medication use, and cardiovascular disease. Model 4 included the same factors in model 3 and additionally adjusted for IOP, equivalent spherical lens, and signal strength index, and excluded patients with a history of DM, hypertension medication use, and cardiovascular disease. Sensistivity analyses were performed by restricting for subjects aged <60 years, healthy subjects, and without carriage of apolipoprotein E4 (APOEε4). For the longitudinal analysis of the COIP cohort, the population was divided into four quartiles (Q1–Q4) by age, SBP, DBP, MAP, and MPP (high and low). Linear mixed models were used to calculate the longitudinal rate for each pRNFLT and mGCIPLT parameter. These variables were set as continuous variables, while univariate and multivariate linear mixed models were used to estimate the effects of different factors (age, sex, SBP, DBP, MAP, and MPP) on the rate of each parameter. Multivariate linear mixed models were adjusted for age, sex, BMI, smoking status, drinking status, education level, HbA1c level, total cholesterol level, TG level, axial length, and IOP. All statistical analyses were performed using STATA software (version 17.0). A two-sided test with a p value of <0.05 was considered significant.

## Results

### Baseline characteristics of the participants in both cohorts

In the UKB, 22,801 participants were included in the study (**Figure 2**). **Table 1** presents the demographic characteristics of the included participants in UKB and COIP. For UKB subjects, the mean age was 55.06 ± 8.26 years, the mean BMI was 27.19 ± 4.67 kg/m^2^, the mean SBP was 135.61 ± 17.95 mmHg, the mean DBP was 81.53 ± 9.94 mmHg, the MAP was 99.56 ± 11.67 mmHg, and the MPP was 54.08 ± 12.90 mmHg. In the COIP cohort, the mean age at baseline was 63.05 ± 8.34 years, the mean SBP was 130.1 ± 16.8 mmHg, the mean DBP was 69.3 ± 9.5 mmHg, and the mean follow-up time was 2.59 ± 0.66 years. The characteristics of the included and excluded subjects of UKB were shown in **Supplementary Table S1**.

**Table 1.** Characteristics of the included participants in the UKB and COIP cohort.

| Characteristics | Cross-section data | Longitudinal data |
| --- | --- | --- |
|  | UKB (n=22801) | COIP (n=2012) |
| Age, year | 55.06 (8.26) | 63.05 (8.34) |
| Female, % | 12160 (53.33%) | 1237 (61.48) |
| Ethnicity, % | White (91.72%) | Chinese (100%) |
| Current smoker, % | 2156 (9.46%) | 230 (11.43) |
| Systolic blood pressure, mmHg | 135.61 (17.95) | 130.1 (16.8) |
| Diastolic blood pressure, mmHg | 81.53 (9.94) | 69.3 (9.5) |
| Mean arterial pressure, mmHg | 99.56 (11.67) | 89.57 (10.62) |
| Mean pulse pressure, mmHg | 54.08 (12.90) | 60.75 (13.69) |
| Arterial Stiffness Index, per m/s | 9.37 (3.29) | NA |
| Body mass index, kg/m<sup>2</sup> | 27.19 (4.67) | 24.39 (8.42) |
| Waist to hip ratio | 0.87 (0.090) | 0.904 (0.070) |
| Current drinker, % | 21049 (92.32%) | 168 (8.35) |
| Below college/university, % | 14327 (62.83%) | 1718 (85.39) |
| History of diabetes mellitus, % | 819 (3.59%) | 464 (23.06) |
| HbA1c, mmol/mol | 35.48 (5.70) | 48.83 (13.29) |
| Total cholesterol, mmol/L | 5.69 (1.11) | 4.93 (1.04) |
| Triglycerides, mmol/L | 1.66 (0.97) | 2.31 (1.62) |
| High density lipoprotein, mmol/L | 1.48 (0.39) | 1.32 (0.41) |
| Low density lipoprotein, mmol/L | 3.54 (0.84) | 3.08 (0.92) |
| Serum creatinine, µmol/L | 72.94 (14.91) | 69.99 (18.81) |
| Microalbuminuria, mg/mL | 27.74 (114.30) | 2.88 (11.63) |
| C-reactive protein, mg/L | 2.32 (3.94) | 2.18 (5.20) |
| Spherical equivalent, diopter | -0.074 (1.885) | 0.277 (1.899) |
| Axial length, mm | NA | 23.48 (0.90) |
| Intraocular pressure, mmHg | 15.21 (2.89) | 16.06 (2.53) |
| Average mRNFLT, µm | 29.69 (4.26) | NA |
| Average mGCIPLT, µm | 75.00 (5.40) | 71.61 (6.19) |
| Average pRNFLT, µm | NA | 110.91 (12.68) |
| Follow-up, year | 0 | 2.49 (0.66) |

UKB=United Kingdom Biobank; COIP=Chinese Ocular Imaging Project; NA = not available; mRNFLT = macular retinal nerve fiber layer thickness; mGCIPL = ganglion cell-inner plexiform layer thickness; pRNFLT = peripapillary retinal nerve fiber layer thickness.

**Figure 2.**
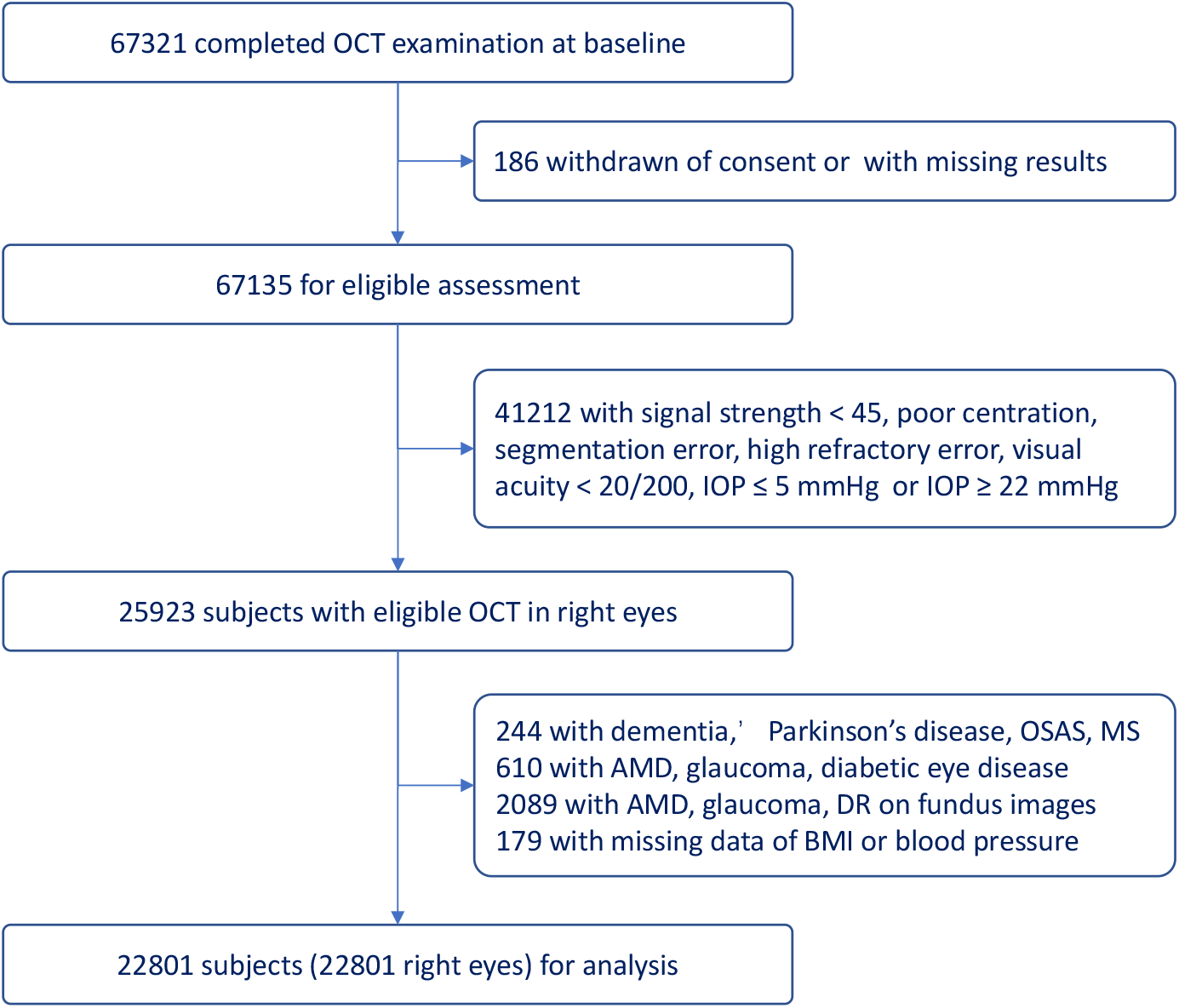
Flow of participants by exclusion from the UK Biobank study.

### Blood pressure and retinal neurodegeneration: cross-sectional data

**Figure 3 and 4** show the mean OCT measurements according to the risk factor quartiles. Each significant risk factor showed strong graded associations with mGCIPLT and mRNFLT measures. After adjusting for the confounders, a thinner mGCIPLT was associated with older age, higher SBP, higher DBP, higher MAP, and higher MPP (**Table 2**). For every 10 mm Hg increase in SBP, the mGCIPL became 0.107-μm thinner (95% confidence interval (CI): 0.066–0.149, p < 0.001). Model 3 and 4 were further corrected for other factors, and the significance of this association was not affected (**Supplementary Table 2**). A thinner mRNFLT was associated with older age, female sex, higher SBP, DBP, MAP, and MPP (**Table 2**). For every 10 mmHg increase in DBP, the mRNFLT became 0.082-μm thinner (95% CI: 0.026–0.138, p = 0.004). Further adjustment largely did not affect this association in model 4. However, no significant relationship was found between ASI and mGCIPLT and mRNFLT. Consistent results were obtained for sensitivity analyses (**Supplementary Table 3-5**).

**Table 2.** Differences in average mGCIPLT and mRNFLT associated with CVD risk factors in UK biobank: cross-sectional data

|  | Model 1 |  | Model 2 |  |
| --- | --- | --- | --- | --- |
| | $\beta$ (95%CI) | P | $\beta$ (95%CI) | P |
| **Average mGCIPLT, $\mu\text{m}$** |  |  |  |  |
| Age, per 10 years | -1.198 (-1.283, -1.114) | <0.001 | -1.205 (-1.290, -1.119) | <0.001 |
| Sex, female | -0.089 (-0.227, 0.049) | 0.209 | -0.068 (-0.208, 0.072) | 0.343 |
| SBP, per 10 mmHg | -0.107 (-0.149, -0.066) | <0.001 | -0.103 (-0.146, -0.061) | <0.001 |
| DBP, per 10 mmHg | -0.197 (-0.268, -0.127) | <0.001 | -0.191 (-0.265, -0.117) | <0.001 |
| MAP, per 10 mmHg | -0.179 (-0.240, -0.117) | <0.001 | -0.174 (-0.238, -0.109) | <0.001 |
| MPP, per 10 mmHg | -0.080 (-0.139, -0.020) | 0.009 | -0.080 (-0.139, -0.020) | 0.009 |
| ASI, per m/s | -0.014 (-0.035, 0.008) | 0.221 | -0.012 (-0.034, 0.010) | 0.275 |
| **Average mRNFLT, $\mu\text{m}$** |  |  |  |  |
| Age, per 10 years | -0.580 (-0.647, -0.513) | <0.001 | -0.582 (-0.650, -0.514) | <0.001 |
| Sex, female | -0.867 (-0.977, -0.758) | <0.001 | -0.846 (-0.957, -0.735) | <0.001 |
| SBP, per 10 mmHg | -0.060 (-0.093, -0.027) | <0.001 | -0.054 (-0.088, -0.020) | 0.002 |
| DBP, per 10 mmHg | -0.082 (-0.138, -0.026) | 0.004 | -0.061 (-0.120, -0.003) | 0.041 |
| MAP, per 10 mmHg | -0.085 (-0.134, -0.037) | 0.001 | -0.071 (-0.122, -0.021) | 0.006 |
| MPP, per 10 mmHg | -0.063 (-0.111, -0.016) | 0.008 | -0.067 (-0.114, -0.019) | 0.006 |
| ASI, per m/s | -0.014 (-0.032, 0.003) | 0.104 | -0.009 (-0.026, 0.009) | 0.316 |

Model 1 adjusting for age, gender, ethnicity, center.
Model 2 adjusting for model 1 plus smoking status, body mass index, alcohol, Townsend deprivation index score, and educational level.
CVD = cardiovascular disease; 95%CI = 95% confidential interval; SBP = systolic blood pressure; DBP = diastolic blood pressure; MAP = mean arterial pressure; MPP = mean pulse pressure; ASI = arterial stiffness index; mRNFLT = macular retinal nerve fiber layer thickness; mGCIPL = ganglion cell-inner plexiform layer thickness.

**Table 3.**
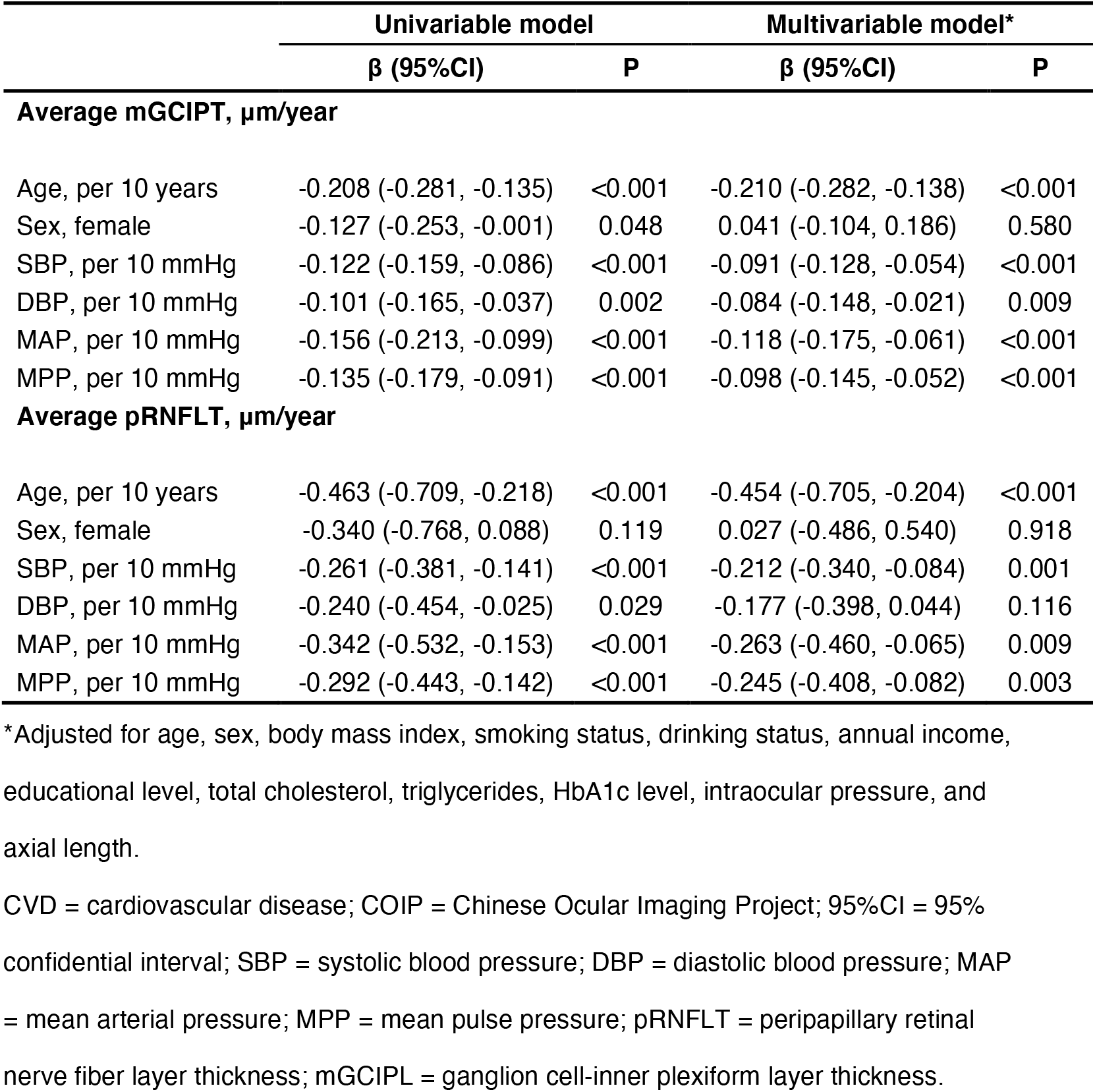
Differences in average mGCIPLT and pRNFLT associated with CVD risk factors in COIP cohort: longitudinal data

**Figure 3.**
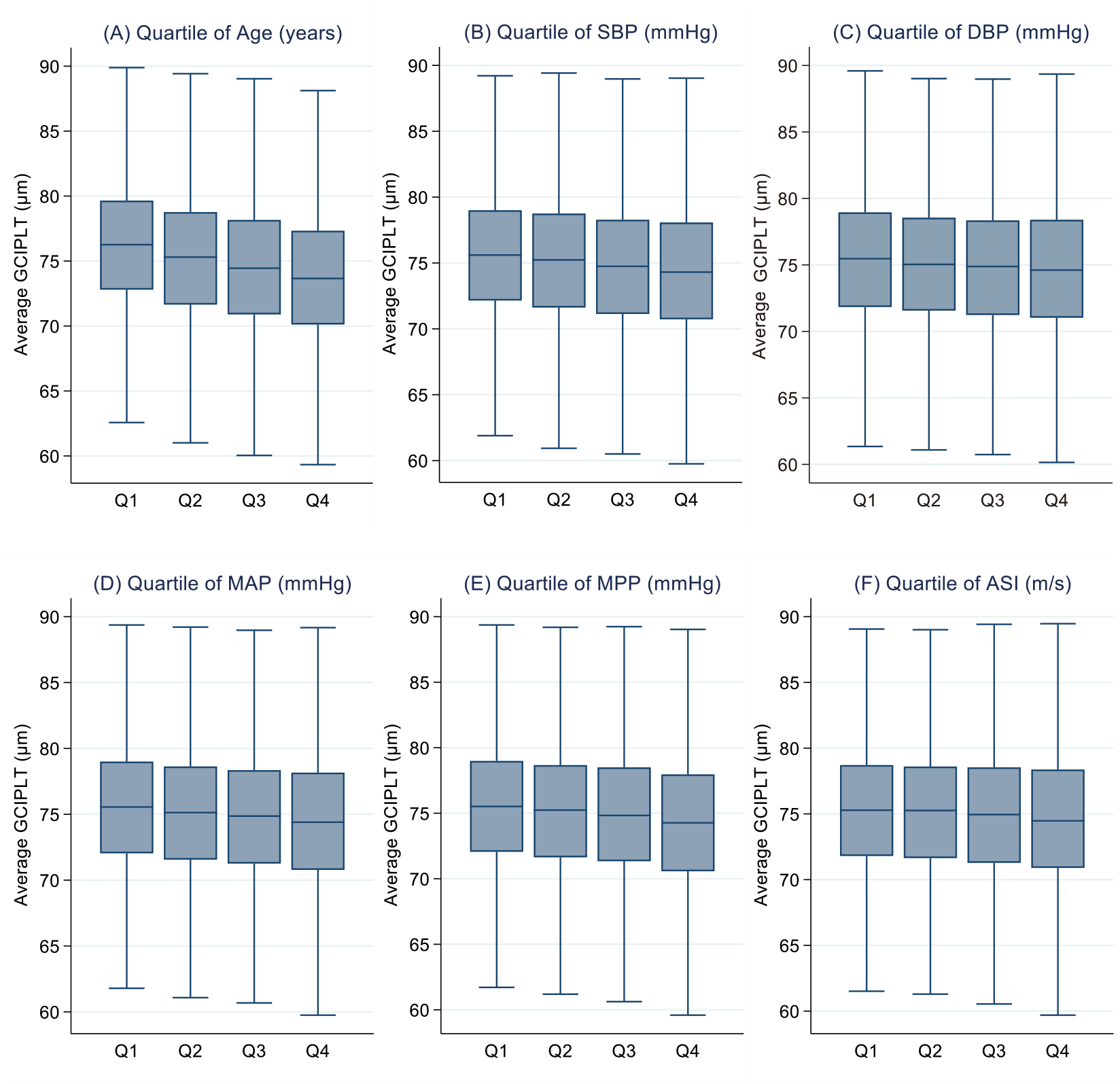
Adjusted mean mGCIPLT by quartiles of cardiovascular disease (CVD) risk factors in UK biobank.

**Figure 4.**
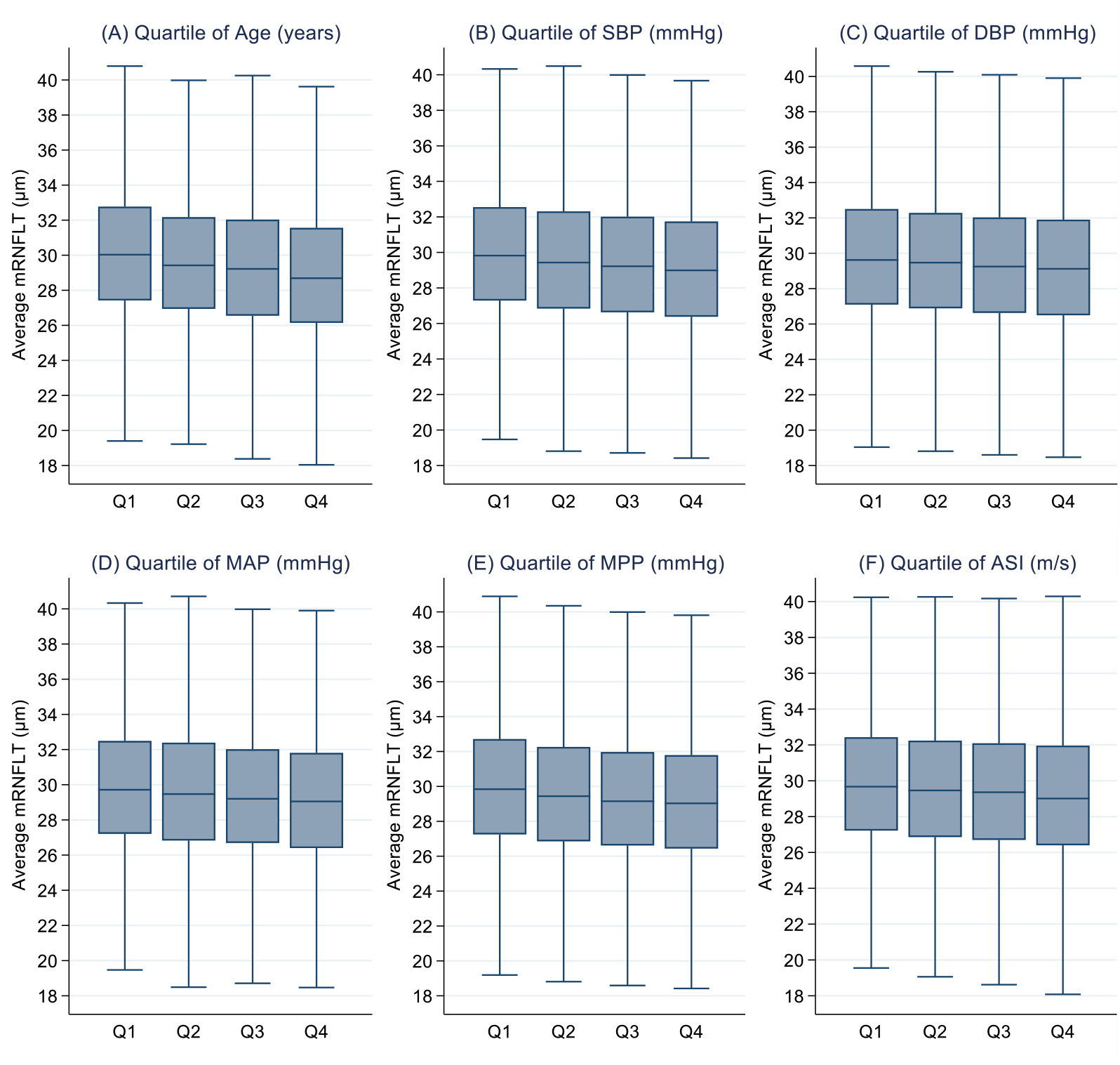
Adjusted mean mRNFLT by quartiles of cardiovascular disease (CVD) risk factors in UK biobank.

### Baseline BP and the slope of retinal neurodegeneration: longitudinal data

**Figure 5** shows the effects of baseline age and BP parameters on the rate of longitudinal changes in mGCIPLT and pRNFLT. The rate of decline in the mean mGCIPLT in different quartiles was significantly affected by age (p for trend < 0.001), SBP (p for trend < 0.001), DBP (p for trend = 0.026), MAP (p for trend < 0.001), and MPP (p for trend < 0.001). Similarly, the rate of decline in the mean mRNFLT was significantly affected by age (p for trend = 0.004), SBP (p for trend < 0.001), DBP (P for trend = 0.014), MAP (p for trend < 0.001), and MPP (p for trend < 0.001). The analysis was performed according to the different subregions, and consistent results were obtained (**Supplementary Table S6-7**).

**Figure 5.**
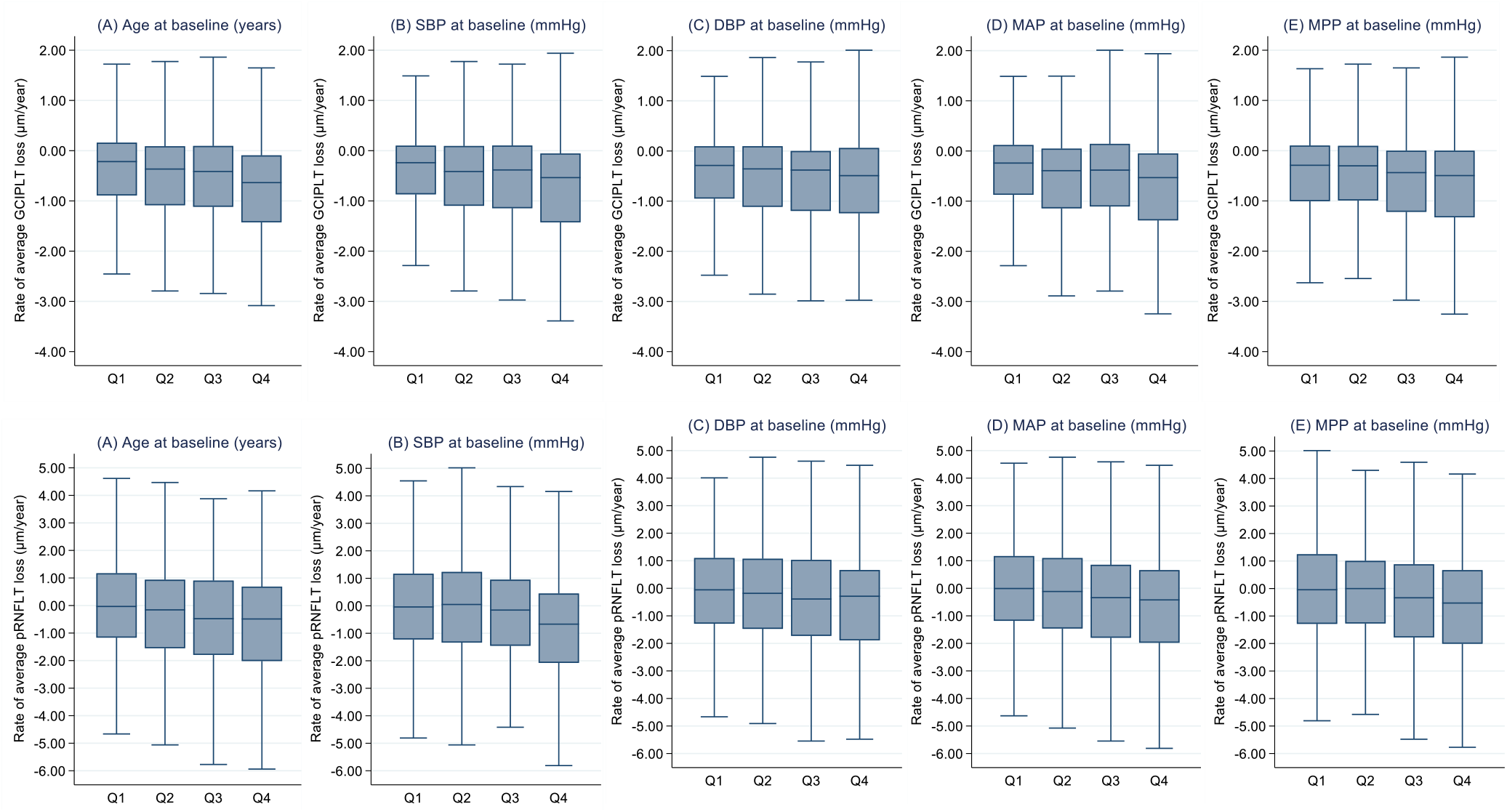
Rate of pRNFLT and mGCIPLT changes overtime by quartiles of cardiovascular disease (CVD) risk factors in COIP cohort.

**Table 3** shows the relationship between BP and the longitudinal rates of retinal neurodegeneration. After adjusting for other factors, accelerated mGCIPLT thinning was associated with older age (β = −0.210, 95% CI: −0.282 to −0.138, p < 0.001), higher SBP (β = −0.091, 95% CI: −0.128 to −0.054, p < 0.001), higher DBP (β = −0.084, 95% CI: −0.148 to −0.021, p = 0.009), higher MAP (β = −0.118, 95% CI: −0.175 to −0.061, p < 0.001), and higher MPP (β = −0.098, 95% CI: −0.145 to −0.052, p < 0.001). Faster pRNFLT thinning was related to older age (β = −0.454, 95% CI: −0.705 to −0.204, p < 0.001), higher SBP (β = −0.212, 95% CI: −0.340 to −0.084, p = 0.001), higher MAP (β = −0.263, 95% CI: −0.460 to −0.065, p = 0.009), and higher MPP (β = −0.245, 95% CI: −0.408 to −0.082, p = 0.001).

## Discussion

Huge strides have been made in the epidemiology, systemic associations, and clinical applications of hypertensive retinopathy over the past three decades. Recent studies have found that optic nerve damage from hypertension is prevalent in the general adult population.^19^ The availability of noninvasive OCT allows the examination of these alterations and their value in hypertension management and dementia risk stratification. This study analyzed the effects of BP and arterial stiffness on RNFLT and GCIPLT, including macular and optic disc area parameters, in two large cohorts. In a cross-sectional study of the UKB, higher SBP, DBP, MAP, and MPP levels were significantly associated with thinner mGCIPLT and mRNFLT, while the ASI level was not significantly associated with mGCIPLT or mRNFLT. Based on the longitudinal data from the Chinese cohort, the higher the SBP, DBP, MAP, and MPP, the faster the longitudinal rate of decline in mGCIPLT. The conclusions are based on sex and diabetes status. Thus, hypertension can not only directly affect the retinal nerve layer thickness but can also accelerate the retinal neurodegeneration with aging.

As the pathology of the retina and brain share a common embryological, anatomical, and immune response basis, the retina has become an effective site for studying the central neurological processes in health and disease.^20^ OCT, a noninvasive and rapid retinal examination, can capture three-dimensional cross-sectional images of the retina and accurately measure the thicknesses of various parts of the retina.^21^ Thinning of the RNFL and GCIPL leads to an increased risk of dementia as well as Alzheimer’s disease, and retinal neurodegeneration can be used as a preclinical biomarker for dementia.

In previous cross-sectional studies, RNFLT was lower in hypertensive patients than in healthy controls. In the European Eye Epidemiology (E3) population, the hypertensive group’s RNFLT was, on average, 0.54-μm thinner than that of the control group.^22^ A longitudinal study by Lee et al. demonstrated that hypertension accelerated the thinning of the pRNFL (hypertensive group: −0.99 μm/y; healthy controls: −0.40 μm/y, p < 0.001).^13^ However, to date, the association between different BP parameters and measurements and RNFLT, which correlates with an accelerated rate of RNFL thinning, has not yet been clarified. The present study is the first to confirm a significant correlation between RNFL thinning and SBP, DBP, MAP, and MPP levels in a cohort with undiagnosed neurodegenerative diseases, such as dementia. Thus, thinning of the retinal nerve layer may be indicative of preclinical central nervous system pathology and is a simple, rapid, and relatively inexpensive predictor of dementia occurrence.

Numerous cross-sectional studies have reported reduced RNFLT levels in patients with dementia. In the Rotterdam Study, each 1 SD decrease in baseline RNFLT was associated with a 44% increase in the risk of dementia.^5^ A 3-year longitudinal cohort study on the UKB also found that changes in RNFLT were associated with cognitive decline, including prospective memory, pairs matching, numeric and verbal reasoning, and reaction time.^6^ In a community-based population cohort of 430 Koreans aged 60 years or older, 215 individuals completed a 5.4-year mean follow-up; participants with a baseline mRNFLT below the lowest quartile cutoff had a greater decline in cognitive scores and a higher prevalence of cognitive impairment and AD compared with those with an RNFLT above the lowest quartile cutoff.^23^ Our study results suggest that monitoring RNFLT in a hypertensive population and using it as a basis for assessing their risk of developing dementia is extremely important. Future studies should assess the predictive strength of RNFLT for the risk of dementia in hypertensive patients to provide more robust evidence for further dissemination of this finding in the clinical setting.

A significant relationship was also observed between SBP, DBP, MAP, MPP, and GCIPL thinning; GCIPL thinning is a possible better marker of dementia.^24–26^ In the Singapore Epidemiology of Eye Diseases Study, the GCIPL was thinner in hypertensive patients than in the healthy population, with varying degrees of thinning across ethnic groups.^27^ For the Rotterdam Study, the risk of dementia was found to be 1.37 per 1 SD reduction in baseline mGCIPL. ^5^A cross-sectional study by Ito et al. showed that the lower region of the mGCIPLT was associated with a 44% increased risk of dementia for every 1 SD decrease in thickness.^28^ Of the total participants, those with lower subcentral concave GCIPLT at baseline had an increased risk of cognitive decline at 3 years (relative risk = 3.49, 95% CI: 1.10–11.1, p = 0.03).^29^ Therefore, monitoring mGCIPLT in hypertensive patients may be useful in assessing their risk of dementia.

Moreover, increased BP levels in DM patients without diabetic retinopathy accelerate the rate of neurodegeneration.^30^ Hypertension may cause thinning of the inner retinal layer, similar to T2DM. This reduction may be associated with damage to the retinal microvasculature from hypertension. Previous studies reported reduced retinal vessel density observed using OCT angiography in hypertensive patients without hypertensive retinopathy; this microvascular damage may affect the atrophy of the inner retinal layer, which is interconnected with blood vessels via the nerves.^14^ Sohn et al. reported that neuronal apoptosis, glial cell reactivity, and many of the abnormal biochemical pathways associated with diabetic neurodegeneration were unrelated to vascular changes, suggesting that neurodegeneration due to DM is nonischemic in nature.^31,32^ Thus, in the presence of DM and hypertension, hypertension-induced ischemic damage leads to more severe inner retinal damage, which accelerates the reduction in the RNFLT and GCIPLT. Microvascular damage related to hypertension onset may accumulate over time and exacerbate the retinal neurodegeneration. However, further studies are needed to confirm this hypothesis.

This study has several implications for public health. Many international guidelines, such as the United States Joint National Committee on Prevention, Detection, Evaluation, and Treatment of High Blood Pressure; the American College of Cardiology/American Heart Association; the European Society of Cardiology/European Society of Hypertension; and the National Institute for Health and Care Excellence (NICE), emphasize the assessment of retinopathy in patients with hypertension via clinical fundus examination or retinography.^33–36^ According to the NICE guidelines, retinal examinations should be offered to all patients with hypertension to assess for hypertensive retinopathy. However, these guidelines have neglected the assessment of retinal neurodegeneration, which can be easily performed, because OCT devices have become commonplace. Evidence suggests that when hypertensive retinopathy is included as an indication for treatment, the percentage of hypertensive patients requiring treatment with antihypertensive medications increases from 3% to 14%. This finding suggests that routine retinal examinations would help physicians consider initiating antihypertensive therapy in patients who are not taking medications or intensifying therapy in patients who are receiving it. Furthermore, the integration of OCT retinal examinations into the evaluation system in the general population without hypertensive emergencies or hypertension may also help with risk stratification and treatment decisions.

These findings have implications for future research. SBP, DBP, MAP, and MPP can influence the extent and rate of retinal neurodegeneration, and should be considered when conducting retinal OCT-related studies. Most OCT studies for eye diseases define controls as individuals without eye disease and rarely exclude people with hypertension or those adjusted for these variables in the statistical analyses. Clinical researchers using OCT to examine RNFLT/GCIPLT in ocular disease should either exclude patients with systemic hypertension from the control group or statistically interpret the systemic vascular risk.^37^ Failure to consider BP factors as potential confounders may bias the study results and lead to erroneous conclusions.

The greatest strength of this study is that it used two large cohorts, the UKB and COIP cohorts, with the largest study sample available, covering the widest range of ethnicities. Second, the parameters measured by OCT were fully automated and quantitative, and prospectively followed up in a longitudinal study that investigated the effect of baseline BP on longitudinal changes in the RNFL and GCIPL, thus revealing a possible causal relationship between BP and retinal nerve layer thinning. Third, the study was adjusted for variables that may affect the assessment of cognitive function (age, sex, education level, diabetes, and APOEε4 status), allowing for a more comprehensive statistical analysis compared with previous investigations.

The present study has some limitations. First, it was not possible to analyze the potential effects of hypertensive medications in detail. A previous study of patients treated for hypertension with either perindopril arginine (an angiotensin-converting enzyme inhibitor) or amlodipine (a calcium channel blocker) found that treatment with perindopril arginine reversed the choroidal thinning (suggesting an improvement and/or reversal of vascular changes), whereas amlodipine use did not significantly affect the choroidal thickness. Whether the inner retinal layer shows a similar response to that of the choroid needs to be investigated. Second, although people with pRNFL defects, glaucomatous neuropathy, and a history of an IOP of ≥21 mmHg were excluded, visual field examinations were not performed; therefore, we cannot completely exclude patients with preclinical glaucoma. Third, we cannot exclude the possibility that the study population included patients with hypertension with a previous occurrence of hypertensive retinopathy that subsequently regressed.

## Conclusion

In summary, a significant relationship was observed between BP metrics and nerve layer thinning in a large population of the UKB and COIP cohorts. An elevated BP level was associated with thinner mRNFL/mGCIPL/pRNFL, even when BP values were below the common treatment thresholds. In DM patients without retinopathy, hypertension exacerbates the rate of retinal neurodegeneration. Understanding the association between BP and RNFL/GCIPL, especially in young adults who have not yet reached the diagnostic threshold for hypertension, may help determine the most effective way to maintain hypertension-related long-term vascular health. These results will broaden the understanding on the relationship between BP and neurodegenerative diseases, predominantly dementia, leading to earlier detection, timely monitoring of neurodegenerative diseases, and improvement in the prognosis of patients with the disease.

## Data Availability

All data produced in the present study are available upon reasonable request to the authors.

## Abbreviations and Acronyms

HbA1c: hemoglobin A1c
BMI: body mass index
SD: standard deviation
SBP: systolic blood pressure
DBP: diastolic blood pressure
MAP: mean arterial pressure
MPP: mean pulse pressure
ASI: arterial stiffness index
CVD: cardiovascular disease
BP: blood pressure
HDL: high density lipoprotein
LDL: low density lipoprotein
mRNFLT: macular retinal nerve fiber layer thickness
mGCIPLT: macular ganglion cell-inner plexiform layer thickness
pRNFLT: peripapillary retinal nerve fiber layer thickness

## Acknowledgements

This research was conducted by the UK Biobank resource under application number 62443. All authors are grateful to the participants of the UK Biobank.

## Personal Thanks

None.

## Funding and Assistance

This study was funded by the National Natural Science Foundation of China (82000901), Fundamental Research Funds of the State Key Laboratory of Ophthalmology (303060202400201209), Guangzhou Science & Technology Plan of Guangdong Pearl River Talents Program (202102010162).

## Author Contributions and Guarantor Statement

W.W. designed the study and performed the statistical analysis. W.W, Z.Z. and M.H. interpreted data. P.Z., G.B. and W.W. interpreted the findings and drafted the manuscript. Z.Z, W.W. and M.H. designed and supervised the study. All authors reviewed the manuscript, edited it for intellectual content, and gave final approval for this version to be published. M.H., Z.Z., and W.W are the guarantors of this work and, as such, had full access to all of the data in the study and take responsibility for the integrity of the data and the accuracy of the data analysis.

## Data and Resource Availability Statement

Data and materials are available via UK Biobank at http://www.ukbiobank.ac.uk/.

## Prior Presentation

This study has not been a prior presentation.

