## Supplementary materials for "Effects of blood pressure and arterial stiffness on retinal neurodegeneration: Cross-sectional and longitudinal evidence from UK Biobank and Chinese cohorts"

**Supplementary Table S1.** Comparisons of the characteristics of the included and excluded subjects in UK biobank.

| **Variable** | **Included** | | | **Excluded** | | | **P-value** |
| --- | --- | --- | --- | --- | --- | --- | --- |
| No. of subjects | 22801 | | | 44334 | | | − |
| Age, year | 55.06 | 8.26 | 57.62 | | 7.96 | <0.001 | |
| Sex, % |  |  |  | |  | <0.001 | |
| Female | 12160 | 53.33% | 24344 | | 54.91% |  | |
| Male | 10641 | 46.67% | 19990 | | 45.09% |  | |
| Ethnicity, % |  |  |  | |  | <0.001 | |
| White | 20912 | 91.72% | 39468 | | 89.02% |  | |
| Eastern Asian | 71 | 0.31% | 237 | | 0.53% |  | |
| Other | 1674 | 7.34% | 4293 | | 9.68% |  | |
| Unknown | 144 | 0.63% | 336 | | 0.76% |  | |
| Smoking status, % |  |  |  | |  | 0.005 | |
| Never | 12588 | 55.21% | 24484 | | 55.23% |  | |
| Former | 7940 | 34.82% | 15202 | | 34.29% |  | |
| Current | 2156 | 9.46% | 4328 | | 9.76% |  | |
| Unknown | 117 | 0.51% | 320 | | 0.72% |  | |
| Townsend deprivation index |  |  |  | |  | <0.001 | |
| Quartile 1 | 4826 | 21.19% | 9110 | | 20.57% |  | |
| Quartile 2 | 5410 | 23.76% | 10003 | | 22.59% |  | |
| Quartile 3 | 6342 | 27.85% | 12235 | | 27.63% |  | |
| Quartile 4 | 6196 | 27.21% | 12932 | | 29.21% |  | |
| SBP, mmHg | 135.61 | 17.95 | 137.91 | | 18.61 | <0.001 | |
| DBP, mmHg | 81.53 | 9.94 | 81.90 | | 10.08 | <0.001 | |
| MAP, mmHg | 99.56 | 11.67 | 100.57 | | 11.87 | <0.001 | |
| MPP, mmHg | 54.08 | 12.90 | 56.01 | | 13.79 | <0.001 | |
| ASI, per m/s | 9.37 | 3.29 | 9.54 | | 4.53 | <0.001 | |
| Body mass index, kg/m^2^ | 27.19 | 4.67 | 27.46 | | 4.83 | <0.001 | |
| Alcohol consumption, % |  |  |  | |  | <0.001 | |
| Never | 917 | 4.02% | 2381 | | 5.37% |  | |
| Previous | 754 | 3.31% | 1640 | | 3.70% |  | |
| Current | 21049 | 92.32% | 40119 | | 90.49% |  | |
| Missing | 81 | 0.36% | 194 | | 0.44% |  | |
| Education attainment, % |  |  |  | |  | <0.001 | |
| College/university or above | 8474 | 37.17% | 15147 | | 34.17% |  | |
| Below college/university | 14327 | 62.83% | 29187 | | 65.83% |  | |
| History of diabetes mellitus, % |  |  |  | |  | <0.001 | |
| No | 21863 | 95.89% | 41122 | | 92.75% |  | |
| Yes | 819 | 3.59% | 2898 | | 6.54% |  | |
| Missing | 119 | 0.52% | 314 | | 0.71% |  | |
| History of CVD |  |  |  | |  | <0.001 | |
| No | 21832 | 95.75% | 41747 | | 94.16% |  | |
| Yes | 969 | 4.25% | 2587 | | 5.84% |  | |
| History of BP treatment |  |  |  | |  | <0.001 | |
| No | 21072 | 92.42% | 39775 | | 89.72% |  | |
| Yes | 1729 | 7.58% | 4559 | | 10.28% |  | |
| HbA1c, mmol/mol | 35.48 | 5.70 | 36.52 | | 7.26 | <0.001 | |
| Total cholesterol, mmol/L | 5.69 | 1.11 | 5.67 | | 1.15 | 0.003 | |
| Triglycerides, mmol/L | 1.66 | 0.97 | 1.68 | | 0.96 | 0.033 | |
| HDL, mmol/L | 1.48 | 0.39 | 1.48 | | 0.39 | 0.495 | |
| LDL, mmol/L | 3.54 | 0.84 | 3.52 | | 0.87 | 0.000 | |
| Spherical equivalent, diopter | -0.074 | 1.885 | -0.517 | | 3.058 | <0.001 | |
| Intraocular pressure, mmHg | 15.21 | 2.89 | 16.42 | | 4.79 | <0.001 | |
| Average mRNFLT, μm | 29.69 | 4.26 | 29.53 | | 10.47 | 0.030 | |
| Average mGCIPLT, μm | 75.00 | 5.40 | 73.02 | | 11.57 | <0.001 | |

SBP = systolic blood pressure; DBP = diastolic blood pressure; MAP = mean arterial pressure; MPP = mean pulse pressure; ASI = arterial stiffness index; CVD = cardiovascular disease; BP = blood pressure; HDL = high density lipoprotein; LDL = low density lipoprotein; mRNFLT = macular retinal nerve fiber layer thickness; mGCIPL = ganglion cell-inner plexiform layer thickness.

**Supplementary Table S2.** Cross-sectional analysis of the differences in average mGCIPLT and mRNFLT associated with CVD risk factors in UK biobank.

| **UK biobank** | **Model 3** | | **Model 4** | |
| --- | --- | --- | --- | --- |
|  | **Coefficient (95%CI)** | **P** | **Coefficient (95%CI)** | **P** |
| **Average mGCIPLT** |  |  |  |  |
| Age, per 10 years | -1.219 (-1.324, -1.114) | <0.001 | -1.570 (-1.675, -1.466) | <0.001 |
| Sex, female | 0.070 (-0.102, 0.242) | 0.426 | 0.150 (-0.017, 0.316) | 0.078 |
| SBP, per 10 mmHg | -0.122 (-0.173, -0.071) | <0.001 | -0.114 (-0.163, -0.064) | <0.001 |
| DBP, per 10 mmHg | -0.214 (-0.303, -0.125) | <0.001 | -0.175 (-0.261, -0.089) | <0.001 |
| MAP, per 10 mmHg | -0.198 (-0.275, -0.121) | <0.001 | -0.173 (-0.248, -0.099) | <0.001 |
| MPP, per 10 mmHg | -0.102 (-0.174, -0.030) | 0.006 | -0.110 (-0.180, -0.040) | 0.002 |
| ASI, per m/s | -0.008 (-0.035, 0.020) | 0.593 | -0.005 (-0.032, 0.022) | 0.707 |
| **Average mRNFLT** |  |  |  |  |
| Age, per 10 years | -0.520 (-0.603, -0.437) | <0.001 | -0.274 (-0.358, -0.189) | <0.001 |
| Sex, female | -0.873 (-1.010, -0.736) | <0.001 | -0.897 (-1.031, -0.763) | <0.001 |
| SBP, per 10 mmHg | -0.069 (-0.110, -0.029) | 0.001 | -0.085 (-0.125, -0.045) | <0.001 |
| DBP, per 10 mmHg | -0.068 (-0.138, 0.003) | 0.060 | -0.105 (-0.174, -0.036) | 0.003 |
| MAP, per 10 mmHg | -0.086 (-0.147, -0.025) | 0.006 | -0.116 (-0.176, -0.056) | <0.001 |
| MPP, per 10 mmHg | -0.094 (-0.152, -0.037) | 0.001 | -0.099 (-0.155, -0.043) | 0.001 |
| ASI, per m/s | -0.005 (-0.027, 0.017) | 0.677 | -0.004 (-0.026, 0.017) | 0.688 |

Model 3 adjusting for age, gender, ethnicity, center, smoking status, body mass index, alcohol consumption, Townsend deprivation index score, educational level, HbA1c, total cholesterol and triglycerides, and excluding those with diabetes, on medication for BP, history of CVD.

Model 4 adjusting for each factor for model 3 plus intraocular pressure, spherical equivalent, signal strength, and excluding those with diabetes, on medication for BP, history of CVD.

CVD = cardiovascular disease; 95%CI = 95% confidential interval; SBP = systolic blood pressure; DBP = diastolic blood pressure; MAP = mean arterial pressure; MPP = mean pulse pressure; ASI = arterial stiffness index; BP = blood pressure; HDL = high density lipoprotein; LDL = low density lipoprotein; mRNFLT = macular retinal nerve fiber layer thickness; mGCIPL = ganglion cell-inner plexiform layer thickness.

**Supplementary Table S3.** Sensitivity analyses by restricting for healthy subjects in UKB

|  | **mGCIPLT** | | **mRNFLT** | |
| --- | --- | --- | --- | --- |
|  | **Coefficient (95%CI)** | **P** | **Coefficient (95%CI)** | **P** |
| **Age, per 10 years** | | | | |
| Model 1 | -1.162 (-1.254, -1.070) | <0.0001 | -0.563 (-0.636, -0.490) | <0.0001 |
| Model 2 | -1.170 (-1.262, -1.077) | <0.0001 | -0.572 (-0.646, -0.498) | <0.0001 |
| Model 3 | -1.195 (-1.307, -1.083) | <0.0001 | -0.520 (-0.609, -0.431) | <0.0001 |
| Model 4 | -1.547 (-1.658, -1.436) | <0.0001 | -0.282 (-0.372, -0.192) | <0.0001 |
| **Sex, female vs male** | | | | |
| Model 1 | -0.038 (-0.188, 0.112) | 0.6184 | -0.879 (-0.998, -0.760) | <0.0001 |
| Model 2 | -0.016 (-0.168, 0.136) | 0.8336 | -0.861 (-0.982, -0.740) | <0.0001 |
| Model 3 | 0.048 (-0.136, 0.231) | 0.6115 | -0.924 (-1.070, -0.778) | <0.0001 |
| Model 4 | 0.126 (-0.051, 0.304) | 0.1624 | -0.950 (-1.093, -0.807) | <0.0001 |
| **SBP, per 10 mmHg** | | | | |
| Model 1 | -0.128 (-0.174, -0.083) | <0.0001 | -0.076 (-0.112, -0.039) | <0.0001 |
| Model 2 | -0.124 (-0.171, -0.078) | <0.0001 | -0.072 (-0.109, -0.035) | 0.0001 |
| Model 3 | -0.134 (-0.189, -0.080) | <0.0001 | -0.080 (-0.124, -0.036) | 0.0003 |
| Model 4 | -0.126 (-0.180, -0.073) | <0.0001 | -0.097 (-0.140, -0.054) | <0.0001 |
| **DBP, per 10 mmHg** | | | | |
| Model 1 | -0.212 (-0.288, -0.135) | <0.0001 | -0.094 (-0.155, -0.033) | 0.0026 |
| Model 2 | -0.210 (-0.290, -0.129) | <0.0001 | -0.079 (-0.143, -0.014) | 0.0167 |
| Model 3 | -0.228 (-0.323, -0.133) | <0.0001 | -0.080 (-0.155, -0.004) | 0.0392 |
| Model 4 | -0.193 (-0.285, -0.100) | <0.0001 | -0.118 (-0.192, -0.043) | 0.0020 |
| **MAP, per 10 mmHg** | | | | |
| Model 1 | -0.200 (-0.267, -0.133) | <0.0001 | -0.102 (-0.156, -0.049) | 0.0002 |
| Model 2 | -0.198 (-0.267, -0.128) | <0.0001 | -0.093 (-0.149, -0.037) | 0.0011 |
| Model 3 | -0.214 (-0.296, -0.132) | <0.0001 | -0.099 (-0.165, -0.034) | 0.0029 |
| Model 4 | -0.191 (-0.271, -0.111) | <0.0001 | -0.132 (-0.196, -0.067) | 0.0001 |
| **MPP, per 10 mmHg** | | | | |
| Model 1 | -0.113 (-0.179, -0.047) | 0.0007 | -0.090 (-0.142, -0.037) | 0.0008 |
| Model 2 | -0.110 (-0.177, -0.044) | 0.0011 | -0.092 (-0.145, -0.040) | 0.0006 |
| Model 3 | -0.119 (-0.197, -0.041) | 0.0029 | -0.109 (-0.171, -0.047) | 0.0006 |
| Model 4 | -0.125 (-0.201, -0.049) | 0.0012 | -0.117 (-0.178, -0.056) | 0.0002 |

Model 1 adjusting for age, gender, ethnicity, center.	Model 2 adjusting for model 1 plus smoking status, BMI, alcohol, Townsend deprivation index score, and educational level. Model 3 adjusting for each factor for model 2 plus HbA1c, total cholesterol and triglycerides and excluding those with diabetes, on medication for BP, history of CVD. Model 4 adjusting for each factor for model 3 plus intraocular pressure, spherical equivalent, signal strength and excluding those with diabetes, on medication for BP, history of CVD.

**Supplementary Table S4.** Sensitivity analyses by restricting for subjects aged < 60 years old in UKB.

|  | **mGCIPLT** | | **mRNFLT** | |
| --- | --- | --- | --- | --- |
|  | **Coefficient (95%CI)** | **P** | **Coefficient (95%CI)** | **P** |
| **Age, per 10 years** | | | | |
| Model 1 | -1.093 (-1.232, -0.954) | <0.0001 | -0.603 (-0.713, -0.493) | <0.0001 |
| Model 2 | -1.108 (-1.249, -0.968) | <0.0001 | -0.610 (-0.721, -0.499) | <0.0001 |
| Model 3 | -1.131 (-1.298, -0.964) | <0.0001 | -0.525 (-0.657, -0.393) | <0.0001 |
| Model 4 | -1.325 (-1.488, -1.163) | <0.0001 | -0.389 (-0.520, -0.258) | <0.0001 |
| **Sex, female vs male** | | | | |
| Model 1 | 0.051 (-0.117, 0.218) | 0.5530 | -0.937 (-1.069, -0.805) | <0.0001 |
| Model 2 | 0.051 (-0.118, 0.221) | 0.5522 | -0.936 (-1.070, -0.802) | <0.0001 |
| Model 3 | 0.224 (0.022, 0.426) | 0.0297 | -0.952 (-1.112, -0.792) | <0.0001 |
| Model 4 | 0.236 (0.041, 0.431) | 0.0179 | -0.932 (-1.089, -0.775) | <0.0001 |
| **SBP, per 10 mmHg** | | | | |
| Model 1 | -0.139 (-0.191, -0.086) | <0.0001 | -0.087 (-0.129, -0.046) | <0.0001 |
| Model 2 | -0.143 (-0.197, -0.089) | <0.0001 | -0.084 (-0.127, -0.041) | 0.0001 |
| Model 3 | -0.156 (-0.220, -0.093) | <0.0001 | -0.093 (-0.143, -0.043) | 0.0003 |
| Model 4 | -0.151 (-0.212, -0.089) | <0.0001 | -0.110 (-0.160, -0.061) | <0.0001 |
| **DBP, per 10 mmHg** | | | | |
| Model 1 | -0.220 (-0.305, -0.135) | <0.0001 | -0.092 (-0.159, -0.025) | 0.0071 |
| Model 2 | -0.234 (-0.323, -0.144) | <0.0001 | -0.084 (-0.155, -0.012) | 0.0214 |
| Model 3 | -0.229 (-0.334, -0.124) | <0.0001 | -0.089 (-0.172, -0.006) | 0.0359 |
| Model 4 | -0.213 (-0.315, -0.112) | <0.0001 | -0.115 (-0.197, -0.033) | 0.0060 |
| **MAP, per 10 mmHg** | | | | |
| Model 1 | -0.208 (-0.283, -0.133) | <0.0001 | -0.107 (-0.166, -0.047) | 0.0004 |
| Model 2 | -0.220 (-0.298, -0.141) | <0.0001 | -0.102 (-0.164, -0.039) | 0.0014 |
| Model 3 | -0.226 (-0.318, -0.134) | <0.0001 | -0.110 (-0.183, -0.037) | 0.0030 |
| Model 4 | -0.215 (-0.304, -0.125) | <0.0001 | -0.136 (-0.208, -0.064) | 0.0002 |
| **MPP, per 10 mmHg** | | | | |
| Model 1 | -0.127 (-0.207, -0.047) | 0.0019 | -0.121 (-0.184, -0.057) | 0.0002 |
| Model 2 | -0.129 (-0.210, -0.049) | 0.0016 | -0.119 (-0.182, -0.055) | 0.0003 |
| Model 3 | -0.163 (-0.258, -0.068) | 0.0007 | -0.135 (-0.210, -0.060) | 0.0004 |
| Model 4 | -0.161 (-0.253, -0.070) | 0.0006 | -0.152 (-0.225, -0.078) | 0.0001 |

Model 1 adjusting for age, gender, ethnicity, center.	Model 2 adjusting for model 1 plus smoking status, BMI, alcohol, Townsend deprivation index score, and educational level. Model 3 adjusting for each factor for model 2 plus HbA1c, total cholesterol and triglycerides and excluding those with diabetes, on medication for BP, history of CVD. Model 4 adjusting for each factor for model 3 plus intraocular pressure, spherical equivalent, signal strength and excluding those with diabetes, on medication for BP, history of CVD.

**Supplementary Table S5.** Sensitivity analyses by restricting for subjects without apolipoprotein E4 (APOEε4) in UK biobank.

|  | **mGCIPLT** | | **mRNFLT** | |
| --- | --- | --- | --- | --- |
|  | **Coefficient (95%CI)** | **P** | **Coefficient (95%CI)** | **P** |
| **Age, per 10 years** | | | | |
| Model 1 | -1.226 (-1.325, -1.127) | <0.0001 | -0.592 (-0.671, -0.514) | <0.0001 |
| Model 2 | -1.230 (-1.330, -1.129) | <0.0001 | -0.595 (-0.675, -0.516) | <0.0001 |
| Model 3 | -1.221 (-1.343, -1.100) | <0.0001 | -0.517 (-0.613, -0.420) | <0.0001 |
| Model 4 | -1.555 (-1.676, -1.434) | <0.0001 | -0.267 (-0.365, -0.170) | <0.0001 |
| **Sex, female vs male** | | | | |
| Model 1 | -0.073 (-0.235, 0.089) | 0.3772 | -0.874 (-1.002, -0.746) | <0.0001 |
| Model 2 | -0.053 (-0.217, 0.111) | 0.5247 | -0.852 (-0.982, -0.722) | <0.0001 |
| Model 3 | 0.033 (-0.165, 0.232) | 0.7415 | -0.876 (-1.034, -0.719) | <0.0001 |
| Model 4 | 0.115 (-0.077, 0.307) | 0.2415 | -0.889 (-1.044, -0.735) | <0.0001 |
| **SBP, per 10 mmHg** | | | | |
| Model 1 | -0.111 (-0.160, -0.062) | <0.0001 | -0.074 (-0.113, -0.035) | 0.0002 |
| Model 2 | -0.106 (-0.156, -0.056) | <0.0001 | -0.069 (-0.109, -0.030) | 0.0006 |
| Model 3 | -0.118 (-0.177, -0.058) | 0.0001 | -0.093 (-0.140, -0.046) | 0.0001 |
| Model 4 | -0.100 (-0.158, -0.043) | 0.0007 | -0.109 (-0.156, -0.063) | <0.0001 |
| **DBP, per 10 mmHg** | | | | |
| Model 1 | -0.194 (-0.277, -0.112) | <0.0001 | -0.105 (-0.170, -0.039) | 0.0018 |
| Model 2 | -0.180 (-0.267, -0.094) | <0.0001 | -0.087 (-0.156, -0.019) | 0.0128 |
| Model 3 | -0.204 (-0.306, -0.101) | 0.0001 | -0.107 (-0.188, -0.025) | 0.0102 |
| Model 4 | -0.154 (-0.254, -0.055) | 0.0024 | -0.143 (-0.224, -0.063) | 0.0005 |
| **MAP, per 10 mmHg** | | | | |
| Model 1 | -0.180 (-0.252, -0.108) | <0.0001 | -0.107 (-0.164, -0.049) | 0.0003 |
| Model 2 | -0.170 (-0.245, -0.095) | <0.0001 | -0.096 (-0.156, -0.036) | 0.0016 |
| Model 3 | -0.190 (-0.279, -0.101) | <0.0001 | -0.123 (-0.193, -0.052) | 0.0006 |
| Model 4 | -0.153 (-0.239, -0.066) | 0.0005 | -0.154 (-0.223, -0.084) | <0.0001 |
| **MPP, per 10 mmHg** | | | | |
| Model 1 | -0.088 (-0.158, -0.018) | 0.0142 | -0.075 (-0.131, -0.020) | 0.0079 |
| Model 2 | -0.090 (-0.160, -0.019) | 0.0128 | -0.080 (-0.136, -0.024) | 0.0053 |
| Model 3 | -0.100 (-0.184, -0.016) | 0.0198 | -0.115 (-0.182, -0.048) | 0.0007 |
| Model 4 | -0.096 (-0.178, -0.015) | 0.0202 | -0.121 (-0.187, -0.056) | 0.0003 |

**Supplementary Table S6.** Average rate of mGCIPLT and pRNFLT changes overtime in COIP cohort during the study period.

|  | **Quartile 1** | **Quartile 2** | **Quartile 3** | **Quartile 4** | **P trend** |
| --- | --- | --- | --- | --- | --- |
| **Average GCIPLT, μm/year** | | | | | |
| Age | -0.388±1.084 | -0.544±1.247 | -0.571±1.635 | -0.838±1.580 | <0.001 |
| SBP | -0.351±1.120 | -0.548±1.363 | -0.582±1.537 | -0.824±1.535 | <0.001 |
| DBP | -0.473±1.392 | -0.516±1.517 | -0.676±1.376 | -0.626±1.297 | 0.026 |
| MAP | -0.356±1.319 | -0.549±1.237 | -0.618±1.657 | -0.768±1.338 | <0.001 |
| MPP | -0.435±1.197 | -0.462±1.127 | -0.626±1.617 | -0.778±1.600 | <0.001 |
| **Average pRNFLT, μm/year** | | | | | |
| Age | -0.141±3.056 | -0.410±3.693 | -0.626±4.224 | -1.040±5.275 | 0.004 |
| SBP | -0.208±3.718 | 0.161±3.546 | -0.670±4.266 | -1.325±4.319 | <0.001 |
| DBP | -0.216±3.877 | -0.447±4.841 | -0.535±3.821 | -0.816±3.220 | 0.048 |
| MAP | -0.093±3.997 | -0.090±3.001 | -0.916±5.259 | -0.916±3.282 | <0.001 |
| MPP | -0.028±3.545 | -0.251±3.564 | -0.829±4.456 | -0.976±4.376 | <0.001 |

COIP = Chinese Ocular Imaging Project; SBP = systolic blood pressure; DBP = diastolic blood pressure; MAP = mean arterial pressure; MPP = mean pulse pressure; pRNFLT = peripapillary retinal nerve fiber layer thickness; mGCIPL = ganglion cell-inner plexiform layer thickness.

**Supplementary Table S7.** Subregional analyzing the rates of mGCIPLT and pRNFLT changes overtime in COIP cohort during the study period.

|  | **mGCIPLT** | | | | **pRNFLT** | | | |
| --- | --- | --- | --- | --- | --- | --- | --- | --- |
|  | **Superior** | **Inferior** | **Nasal** | **Temporal** | **Superior** | **Inferior** | **Nasal** | **Temporal** |
| **Age** |  |  |  |  |  |  |  |  |
| Quartile 1 | -0.114±0.649 | -0.150±0.657 | -0.659±0.950 | 0.092±0.733 | -1.903±6.078 | -0.475±6.490 | -0.5028.911 | 2.363±9.206 |
| Quartile 2 | -0.214±1.367 | -0.191±1.061 | -0.798±1.166 | -0.009±2.088 | -1.163±7.347 | -1.179±7.205 | -0.678±8.268 | 1.387±6.915 |
| Quartile 3 | -0.189±1.681 | -0.250±1.568 | -1.120±2.220 | 0.115±2.009 | -1.045±7.902 | -1.361±7.974 | -1.470±10.407 | 1.373±7.833 |
| Quartile 4 | -0.269±1.725 | -0.324±1.361 | -1.084±1.601 | -0.002±1.409 | -1.118±8.831 | -2.166±8.954 | -1.742±9.902 | 0.961±7.394 |
| P for trend | 0.261 | 0.087 | <0.001 | 0.787 | 0.136 | 0.006 | 0.050 | 0.028 |
| **SBP** |  |  |  |  |  |  |  |  |
| Quartile 1 | -0.139±0.857 | -0.189±0.817 | -0.759±1.235 | 0.088±1.009 | -2.007±6.766 | -0.552±7.850 | -0.541±8.752 | 2.331±7.381 |
| Quartile 2 | -0.226±1.135 | -0.100±0.971 | -0.910±1.092 | 0.115±1.633 | -0.518±6.092 | 0.109±7.518 | -0.585±9.478 | 1.656±9.112 |
| Quartile 3 | -0.162±2.024 | -0.259±1.748 | -0.933±2.223 | 0.196±2.086 | -0.935±8.003 | -1.767±7.426 | -1.274±9.944 | 1.349±6.937 |
| Quartile 4 | -0.244±1.364 | -0.365±1.071 | -1.024±1.527 | -0.252±1.904 | -1.822±8.686 | -2.676±7.139 | -1.739±9.171 | 0.927±8.190 |
| P for trend | 0.497 | 0.053 | 0.046 | 0.080 | 0.840 | <0.001 | 0.053 | 0.015 |
| **DBP** |  |  |  |  |  |  |  |  |
| Quartile 1 | -0.198±1.502 | -0.249±1.057 | -0.805±1.282 | 0.052±1.846 | -1.115±6.581 | -0.971±7.792 | -0.756±8.065 | 1.976±7.429 |
| Quartile 2 | -0.137±1.280 | -0.135±0.950 | -0.835±1.293 | 0.195±1.863 | -1.679±7.977 | -0.905±8.912 | -1.292±10.331 | 2.193±6.607 |
| Quartile 3 | -0.291±1.670 | -0.289±1.680 | -0.963±1.346 | -0.095±1.638 | -1.287±7.745 | -1.160±7.070 | -1.098±9.793 | 1.422±8.530 |
| Quartile 4 | -0.124±0.833 | -0.202±0.894 | -0.992±2.234 | 0.041±1.002 | -1.237±7.429 | -1.787±6.072 | -0.919±8.994 | 0.687±9.060 |
| P for trend | 0.908 | 0.955 | 0.101 | 0.459 | 0.990 | 0.135 | 0.890 | 0.014 |
| **MAP** |  |  |  |  |  |  |  |  |
| Quartile 1 | -0.098±0.910 | -0.174±0.889 | -0.722±1.207 | 0.202±1.489 | -1.502±6.805 | -0.435±8.100 | -1.013±9.336 | 2.642±7.886 |
| Quartile 2 | -0.228±1.529 | -0.214±0.955 | -0.926±1.345 | 0.044±1.623 | -0.976±6.155 | -0.707±6.649 | -0.265±7.576 | 1.611±6.006 |
| Quartile 3 | -0.215±1.710 | -0.157±1.730 | -0.918±1.377 | 0.070±1.721 | -1.480±8.955 | -1.484±8.704 | -1.884±10.808 | 1.225±8.871 |
| Quartile 4 | -0.233±1.295 | -0.347±1.008 | -1.053±2.211 | -0.155±1.826 | -1.356±7.631 | -2.197±6.416 | -0.900±9.281 | 0.796±8.582 |
| P for trend | 0.250 | 0.172 | 0.015 | 0.019 | 0.992 | 0.001 | 0.578 | 0.001 |
| **MPP** |  |  |  |  |  |  |  |  |
| Quartile 1 | -0.132±0.742 | -0.168±0.690 | -0.740±1.102 | 0.047±0.962 | -1.525±6.559 | -0.356±7.468 | -0.033±9.600 | 1.872±8.879 |
| Quartile 2 | -0.164±0.885 | -0.113±0.914 | -0.813±1.042 | 0.127±1.496 | -1.255±6.397 | -0.438±7.395 | -1.195±7.988 | 1.892±7.227 |
| Quartile 3 | -0.225±2.092 | -0.264±1.853 | -1.165±2.402 | 0.183±2.228 | -1.182±8.671 | -1.794±7.410 | -1.710±10.487 | 1.401±8.095 |
| Quartile 4 | -0.259±1.605 | -0.374±1.111 | -0.915±1.438 | -0.173±1.917 | -1.341±8.131 | -2.368±7.825 | -1.305±9.060 | 1.123±7.346 |
| P for trend | 0.234 | 0.024 | 0.023 | 0.274 | 0.702 | <0.001 | 0.040 | 0.146 |

COIP = Chinese Ocular Imaging Project; SBP = systolic blood pressure; DBP = diastolic blood pressure; MAP = mean arterial pressure; MPP = mean pulse pressure; mGCIPL = ganglion cell-inner plexiform layer thickness; pRNFLT = peripapillary retinal nerve fiber layer thickness.
